# The effect of community-based behaviour change interventions on observed handwashing and child faeces disposal in rural Malawi: a controlled before-and-after (CBA) trial

**DOI:** 10.1101/2025.06.12.25329256

**Authors:** Clara MacLeod, Mindy Panulo, Joseph Wells, Blessings White, Ian Ross, Tracy Morse, Robert Dreibelbis, Kondwani Chidziwisano

**Author notes:** These authors contributed equally to the work (Robert Dreibelbis, Kondwani Chidziwisano). Corresponding author: Clara MacLeod, London School of Hygiene and Tropical Medicine, Keppel Street, London WC1E 7HT United Kingdom. **Trial registration**: The study was registered at clinicaltrials.gov (NCT05808218); https://clinicaltrials.gov/study/NCT05808218.

## Abstract

**Introduction:** While hygiene promotion is frequently included in sanitation programmes, few studies directly evaluate the effect of these interventions on hygiene outcomes. We assessed the effectiveness of a district-level Community-led Total Sanitation (CLTS) intervention with and without locally managed Care Groups (CG) on observed sanitation and hygiene behaviours in a rural area of Chiradzulu District, Malawi.

**Methods:** This was a controlled before-and-after trial with two intervention arms and a control group implemented in three sub-districts (also known as Traditional Authorities [TAs]) in Chiradzulu District, Malawi. Two TAs received CLTS, which included an additional low-intensity government-led hygiene promotion campaign. One TA received the standard CLTS intervention (CLTS group), and one received the same intervention but with additional sanitation and hygiene promotion delivered through local Care Groups (CLTS+CG group). The third TA served as the control group. Hygiene and child faeces disposal outcomes were measured by fieldworker direct observation in a purposively sampled subset of 96 to 140 households per arm enrolled in the main trial at baseline (June 2023) and at endline (May 2024). We estimated intervention effects on observed handwashing behaviour and safe child faeces disposal with hierarchical logistic regression models.

**Results:** In our per protocol analysis, neither intervention was associated with differences in observed handwashing with soap compared to the control arm nor with differences in observed child faeces disposal. Additional analyses found that both interventions were associated with large increases in hand rinsing with water only compared to control groups (CLTS+CG group adjusted relative risk ratio [aRRR] = 2.80, CI = 1.81, 4.33; CLTS group aRRR = 1.76, CI = 1.10, 2.83). The CLTS+CG intervention was associated with a slight increase in hand rinsing compared to the CLTS only group (aRRR = 1.65, CI = 1.01, 2.69).

**Conclusion:** The CLTS+CG and CLTS interventions were associated with an increase in hand rinsing at critical junctures but not handwashing with soap, suggesting that current hygiene programmes require further focus on soap access and use. Future research should investigate barriers to the availability and competing uses of soap in households. Both interventions did not increase safe child faeces disposal, though the sample size was small. Future research should investigate further integration of child faeces management into CLTS and CG behaviour change interventions.

## Introduction

Poor water, sanitation, and hygiene (WASH) is associated with a range of poor health outcomes, such as diarrhoea, acute respiratory infections, undernutrition, and soil-transmitted helminthiasis^1^. Poor WASH also affects educational outcomes, cognitive development, mental wellbeing, and quality of life^2–4^. Improving sanitation coverage and use is associated with reduced exposure to enteric pathogens and reduced diarrhoeal disease^5^, as sanitation facilities serve as a crucial barrier for separating faecal pathogens from the environment. Safe child faeces disposal, often overlooked in sanitation interventions, can also be effective at preventing diarrhoea transmission^6^. Improving handwashing with soap is also associated with reductions in both diarrhoeal disease and acute respiratory infections by over 20%^7,8^, as well as the reduction of certain neglected tropical diseases and some soil-transmitted helminth infections^9,10^.

Behaviour-centred interventions are associated with improved uptake of some sanitation and hygiene practices^11,12^, but there is limited evidence on the effectiveness of integrating hygiene promotion into sanitation interventions on certain sanitation and hygiene outcomes. Community-led Total Sanitation (CLTS) is an approach used globally for promoting household sanitation in rural areas. It centres on community-wide behaviour change and community self-enforcement. Introduced in Bangladesh in 1999 and now adopted by many governments, including the Government of Malawi, CLTS includes a series of community participation activities primarily designed to end open defecation^13^. Evidence on the effectiveness of CLTS in sustaining sanitation gains after ODF achievement is mixed, despite its widespread adoption^14^. CLTS usually incorporates messages to promote handwashing with soap through participatory acitivities^13^, however, these activities are limited in scope. The few studies that evaluated the effect of CLTS on handwashing with soap found no or modest improvements^15–17^. Evidence of CLTS on safe child faeces disposal practices is also limited^6,18^, and few countries include safe child faeces disposal in their requirements for ODF status^19^. In Malawi, supplementing CLTS with additional messaging, such as through government-led handwashing and sanitation promotion campaigns, provides an opportunity to target these behaviours within existing community structures.

The Care Group (CG) model provides an additional opportunity for promoting sanitation and hygiene at the community level^20^. The CG Model has been widely implemented by non-governmental organisations (NGOs) and integrated into national policy in several countries, including Malawi^21^. The CG model aims to reach a high number of households through the development of a supportive network of peer-to-peer counselling for the delivery of health interventions in rural areas^22–26^. The CG model has been effective at improving health and nutrition outcomes^27^. In Malawi, the CG model has primarily focused on nutrition interventions^22^. However, there is limited evidence globally on the effectiveness of the CG model for changing WASH behaviours^28,29^ and how the CG model can be integrated into existing WASH interventions^29^.

To address this gap, we conducted a controlled before-and-after (CBA) trial to assess the effectiveness of two community-based sanitation and hygiene promotion interventions on handwashing and safe child faeces disposal. This study aimed to assess the effectiveness of a CLTS intervention and government-led hygiene promotion with and without the CG model on observed hand hygiene behaviour and safe child faeces disposal in rural Chiradzulu District, Malawi. The objectives were to assess: 1) whether the two interventions were individually more effective than no intervention at all, and 2) how the two interventions compare to each other.

## Methods

This study is embedded within a larger trial that assessed the effect of the same interventions on sanitation coverage and use. Full details of the study design, setting, intervention, and corresponding process evaluation for the larger trial are reported elsewhere^30,31^. Key study information is summarised briefly below, as well as additional details on the hygiene promotion components of the intervention.

### Study design and participants

This study is a CBA trial embedded within the Water, Sanitation, and Hygiene for Everyone (W4E) programme, which targeted universal access to WASH in Chiradzulu District by 2024. The trial had three arms: i) a low-intensity government-led hygiene promotion campaign combined with standard CLTS (henceforth referred to as the CLTS group), ii) a low-intensity government-led hygiene promotion campaign combined with a modified CLTS intervention that incorporated the Care Group model (henceforth referred to as CLTS+CG), and iii) no intervention (henceforth referred to as the control). The control arm received no intervention prior to data collection but received the CLTS intervention after the conclusion of data collection.

### Sample size

The structured observations were nested within the larger W4E trial. See Chidziwisano et al. (2025) details on sample size calculations^31^. In brief, the main study was powered to detect a 13.6% percentage point change in household sanitation coverage, the primary outcome of the trial. We did not conduct formal sample size calculations for this sub-study as structured observational data could only be collected in a purposively selected subset of households included in the main trial. The number of enrolled households ranged 96 to 140 per arm at baseline (June 2023) and endline (May 2024) (Figure 1).

**Figure 1.**
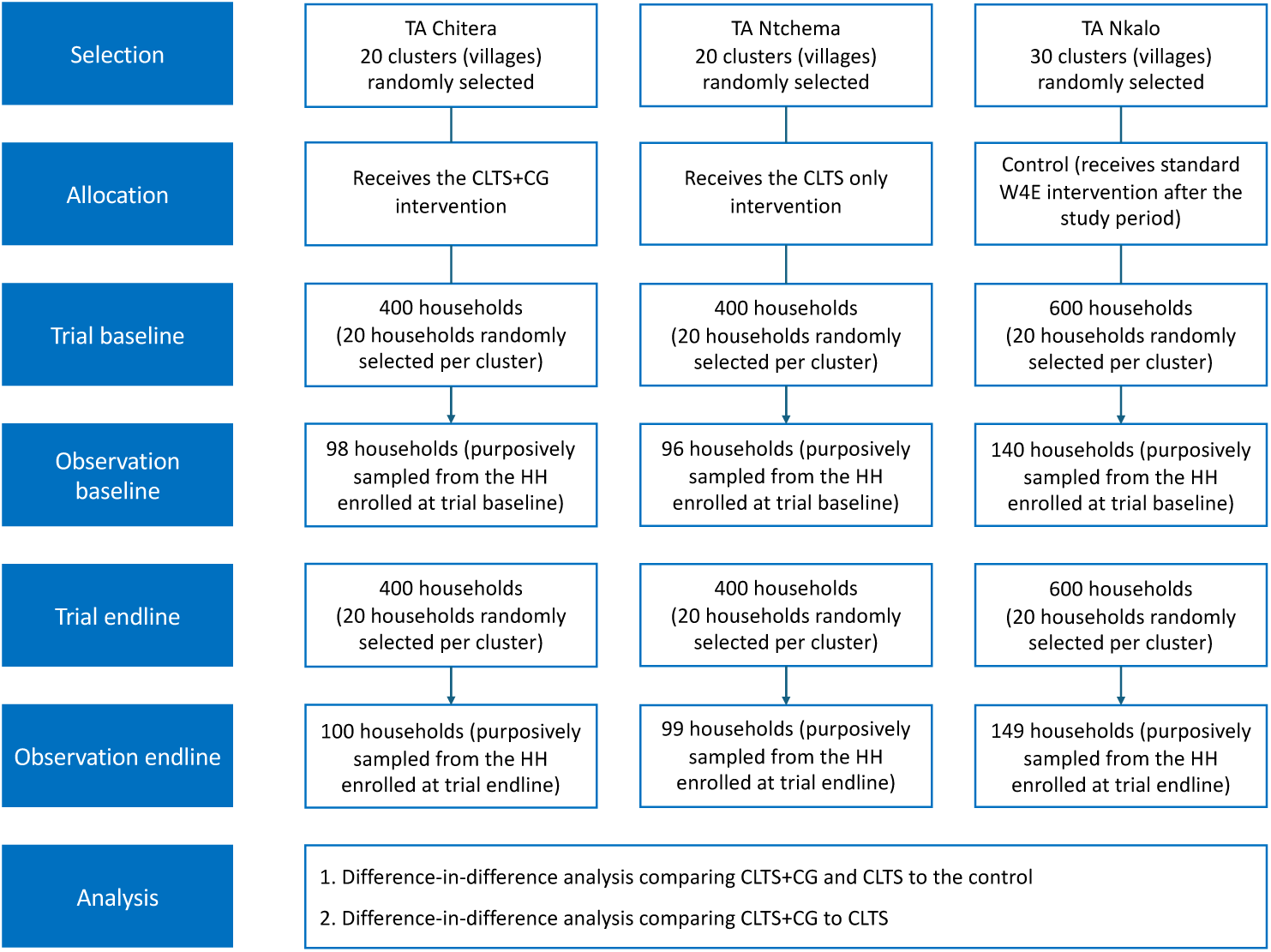
Participant flow diagram.

### Study setting and population

The study took place in Chiradzulu District, Malawi (Figure 4). Chiradzulu District is situated in the southern region of Malawi and is sub-divided into 10 administrative regions known as Traditional Authorities (TAs)^31^. This sub-study was implemented in three TAs: CLTS+CG in Chitera, CLTS in Ntchema, and no intervention in Nkalo (Figure 2). The TAs were the units of allocation.

**Figure 2.**
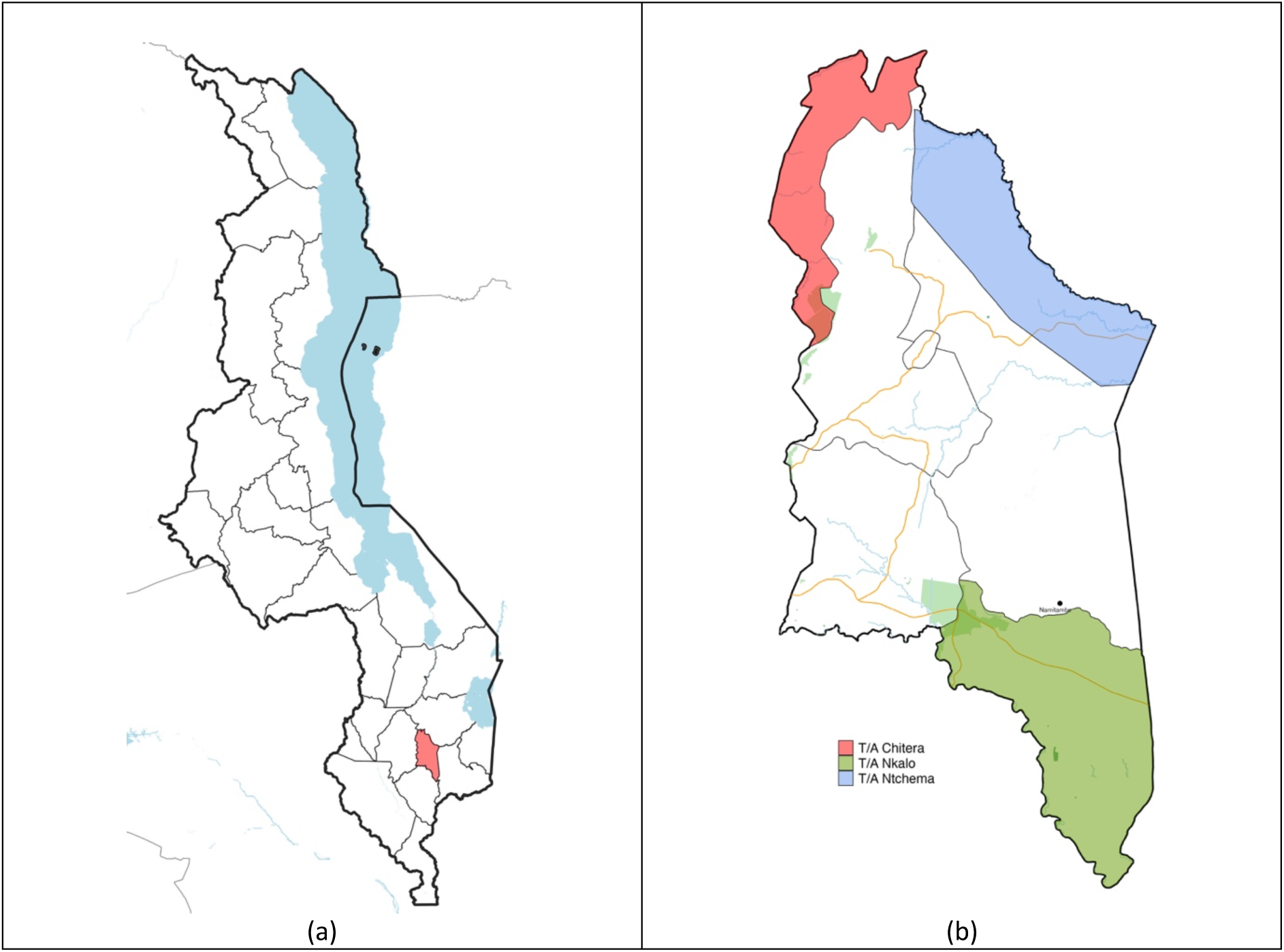
(a) Map of Malawi with Chiradzulu District highlighted in red; (b) Map of Chiradzulu District, Malawi, with TA Chitera (CLTS+CG) highlighted in red, TA Ntchema (CLTS) highlighted in yellow, and TA Nkalo (control) highlighted in green. The base layer was obtained from Database of Global Administrative Areas (GADM) (available from: https://gadm.org/download_country.html). The terms of use can be found here: https://gadm.org/license.html. Road and waterbody features were obtained from OpenStreetMap under the Open Database License (available from: https://www.openstreetmap.org).

### Description of the intervention

Details on the sanitation components of the main intervention are fully elsewhere^31^; additional details on the hygiene promotion activities are described here.

#### CLTS

Both intervention arms received a CLTS intervention, delivered according to the Malawi National Sanitation and Hygiene Strategy^32^ and CLTS Handbook^13^. The CLTS activities included messages delivered during CLTS events to promote sanitation and handwashing with soap through participatory activities. The CLTS programme was augmented with an additional government-led hygiene promotion campaign using pre-established sanitation and hygiene messages on handwashing with soap and safe child faeces disposal from the Ministry of Health (MoH). This was delivered through four separate campaigns each lasting five days. The campaign consisted of a mobile van traveling to marketplaces and village centres to deliver messages via loudspeaker and distribute informational leaflets. There were no household visits or group-based activities.

#### CLTS+CG

In the CLTS+CG arm, households received the CG model, in addition to the CLTS and government-led hygiene promotion activities outlined above. This arm used locally established CGs to deliver messages related to CLTS to households, in addition to those delivered by community health workers (HSAs) and natural leaders. Specifically, the CGs conducted household CLTS follow-up visits to households in their communities to track progress against sanitation coverage and provide additional messages on sanitation and hygiene.

#### Control

TA Nkalo, the control arm, did not receive the CLTS and government-led hygiene promotion intervention during the study period.

### Selection of clusters and households

Clusters were the communities (villages) where interventions were delivered. The selection of households and clusters for the full trial is described elsewhere^31^. For this sub-study, a subset of households was purposively selected for structured observations on handwashing and safe child faeces disposal. 334 households (98 in the CLTS+CG group, 96 in the CLTS group, and 140 in the control group) were selected for direct observation of sanitation and hygiene behaviours at baseline. At endline, 348 households were selected (100 in the CLTS+CG group, 99 in the CLTS group, and 149 in the control group). Households were purposively selected from households participating in the household survey with a child under the age of five.

### Data collection

We conducted the baseline data collection between April and May 2023 in the two treatment arms prior to the implementation of the W4E interventions. The eleven-month follow-up survey was conducted in March and April 2024 before World Vision and Water for People implemented CLTS in the control arm (TA Nkalo). A structured questionnaire with close-ended questions and pre-coded responses was used to collect data on household characteristics and sanitation and hygiene outcomes. Trained enumerators conducted the household survey in Chichewa, the main language spoken in Chiradzulu District, and recorded answers using mobile devices using Kobo Collect software. The trained enumerators also conducted structured observations in a purposively selected sample of households randomly selected to participate in the larger trial^31^. The structured observations captured handwashing behaviour at key pre-defined junctures and child faeces disposal. Trained enumerators visited a household between 8:00am to 12:00 noon, as behaviours of interest mostly occur in the morning hours to noon^33^. Observations lasted up to 4 hours.

### Outcomes

The first outcome was whether or not observed hand hygiene opportunities were accompanied by hand handwashing with soap (Table 1). Hand hygiene opportunities were defined as before food preparation, before eating, before child feeding, after using a latrine, after cleaning a child, and after coming into contact with an animal. The second outcome, safe child faeces disposal is defined according to the WHO/UNICEF Joint Monitoring Programme for Water Supply, Sanitation, and Hygiene (JMP) definition^34^ (Table 1). During the analysis stage, we specified an additional hand hygiene outcome, a three-level categorical variable that includes no hand hygiene action, hand rinsing, and handwashing with soap at the same pre-specified junctures (Table 1).

**Table 1.** Description of study outcomes.

| Outcomes | Description |
| --- | --- |
| Handwashing with soap | Binary outcome based on observed hand hygiene opportunities (i.e., before food preparation, before eating, before child feeding, after using a latrine, after cleaning a child, and after coming into contact with an animal) accompanied by handwashing with soap and water compared to no action. |
| Safe child faeces disposal | Binary outcome based on observed household disposal site of last defaecation or child latrine use for all children under the age of five years. Safe disposal methods and sites include putting or rinsing the child faeces into the latrine or toilet, disposing into the garbage, or burying. |
| Hand hygiene behaviour | A three-level categorical variable based on observed hand hygiene opportunities (i.e., before food preparation, before eating, before child feeding, after using a latrine, after cleaning a child, and after coming into contact with an animal) accompanied by either: i) handwashing with soap, ii) hand rinsing (washing hands with water only), or iii) no action. |

### Data analysis

Data was cleaned using R statistical software^35^ and analysed using Stata v18 (Stata Corp, College Station, TX). We used an intention-to-treat approach for analysis where we analysed data according to treatment arm assignment^36^. A difference-in-difference (DID) analysis using regression approaches was used to estimate the intervention effect adjusted for baseline differences in observed handwashing behaviour and safe child faeces disposal. We produced measures of association with two sets of regression models which were developed for all outcomes. In the first set of models, we included design variables only (intervention assignment and community size). The second set of models were adjusted for pre-specified covariates, including number of household members, at least one household member with a disability, wealth quintile based on HH assets^37^, water source on plot, and village size.

For our two pre-specified binary outcomes, DID models were estimated using multilevel mixed-effects logistic regression with robust standard errors to account for clustering at the village level (*melogit*)^38–40^. Regression coefficients were converted to odds ratios. For each behavioural outcome, we reported the odds ratio, 95% confidence interval, and p-value for both treatment arms compared to the control, as well as for the CLTS+CG group compared to the CLTS group.

We conducted an additional analysis on a three-level outcome variable for observed hand hygiene behaviour (i.e., handwashing with soap, hand rinsing, or no action) using multinomial logistic regression accounting for clustering at the village level (*mlogit*). For this behavioural outcome, we reported the relative risk ratio (RRR), 95% confidence interval, and p-value for both treatment arms compared to the control, as well as for the CLTS+CG group compared to the CLTS group.

### Ethics

Ethical approval was obtained from the Malawi National Commission for Science and Technology, Lilongwe, Malawi (reference number P01/23/718) and the London School of Hygiene and Tropical Medicine Research Ethics Committee, London, United Kingdom (reference number 30722). Participants were provided with an information sheet explaining the study procedure and aim prior to study activities. Informed written consent was obtained from all study participants recruited into the study prior to data collection. The study was registered at clinicaltrials.gov (NCT05808218).

## Results

### Household characteristics

We conducted direct observations in 334 households at baseline and 348 households at endline (Table 2). The median household size was five members. According to the Washington Group short form^41^, 3% of households had at least one member living with a disability at baseline and 5% at endline. Two percent of households had a water source on their plot at baseline and 5% at endline. Over half of households had an observed or reported handwashing facility at baseline (60%) and endline (54%). Soap was observed at only 8% of handwashing facilities at both baseline and endline.

**Table 2.**
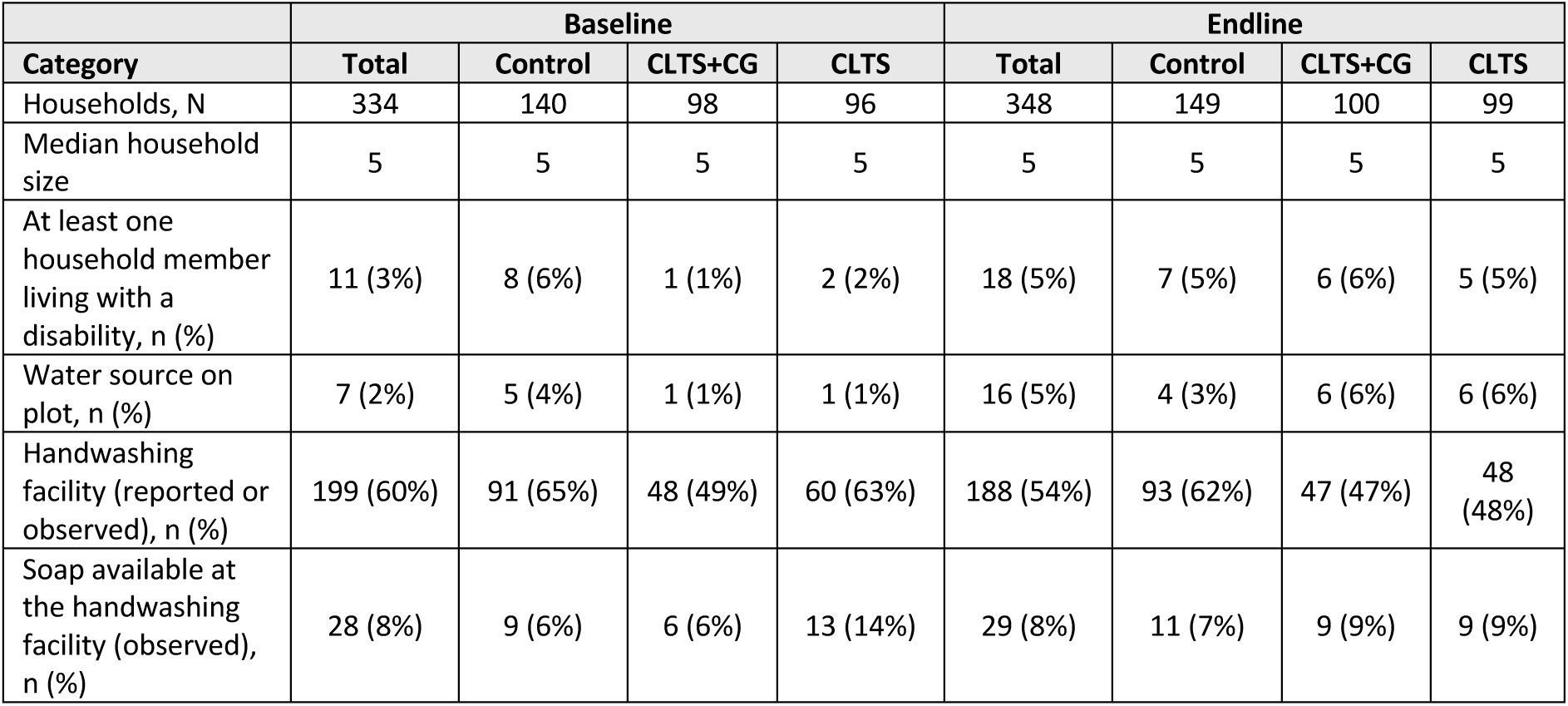
Characteristics of households enrolled in the study at baseline and endline.

### Observed hand hygiene behaviour

We observed 1,099 opportunimes for handwashing across all intervenmon groups at baseline and 1,310 opportunimes at endline (Figure 3, Table 3). Per our pre-specified analysis, we found no evidence of differences in the odds of handwashing with soap among households in the CLTS group (adjusted odds ratio [aOR] = 0.75, 95% confidence interval [CI] = 0.13, 4.33) and CLTS+CG group (aOR = 1.87, CI = 0.54, 6.40) compared to households in the control group eleven months after the intervention (Table S1). We also found no evidence of a difference in the odds of handwashing with soap in the CLTS+CG group compared to the CLTS group (aOR = 2.78; CI = 0.49, 15.64) (Table S2).

**Figure 3.**
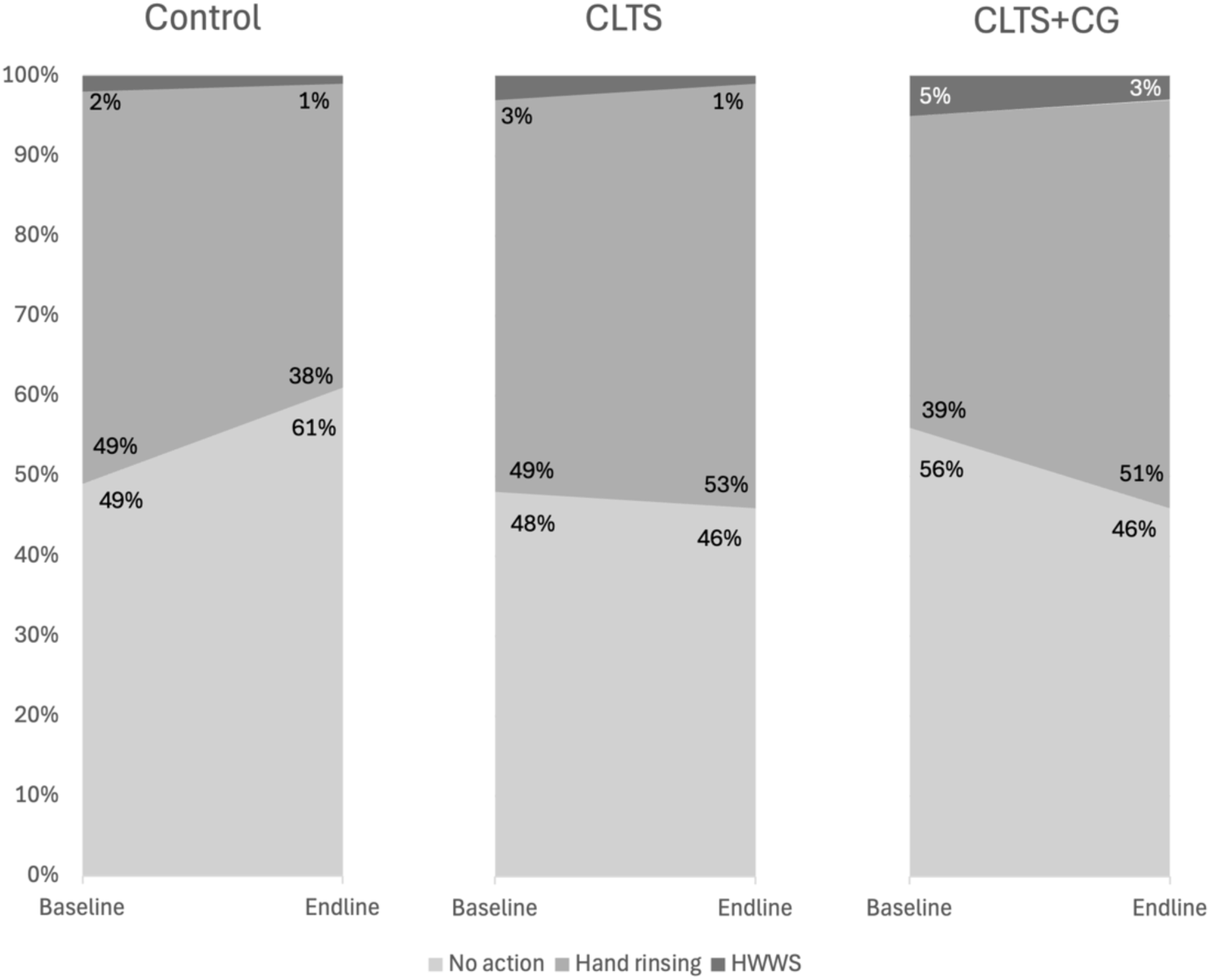
Observed hand hygiene behaviour at baseline and endline in each trial arm. HWWS = handwashing with soap.

**Table 3.** CLTS and CLTS+CG vs. control for the three-level handwashing outcome variable.

|  | Baseline, n (%) | Endline, n (%) | Difference, % | RRR (95% CI) | aRRR (95% CI) |
| --- | --- | --- | --- | --- | --- |
| **Handwashing with soap and water** |  |  |  |  |  |
| CLTS+CG | 17 (5%) | 12 (3%) | -2 | 2.19 (0.65, 7.37) | 3.11 (0.91, 10.59) |
| CLTS | 7 (3%) | 3 (1%) | -1 | 1.16 (0.20, 7.09) | 1.08 (0.18, 6.47) |
| Control | 12 (2%) | 7 (1%) | -2 | 1.00 (Ref) | 1.00 (Ref) |
| **Hand rinsing** |  |  |  |  |  |
| CLTS+CG | 128 (39%) | 181 (51%) | 11 | 2.60 (1.75, 3.86) | 2.80 (1.81, 4.33) |
| CLTS | 146 (49%) | 169 (53%) | 2 | 1.81 (1.12, 2.94) | 1.76 (1.10, 2.83) |
| Control | 231 (49%) | 244 (38%) | -12 | 1.00 (Ref) | 1.00 (Ref) |
| **No action** |  |  |  |  |  |
| CLTS+CG | 185 (56%) | 161 (46%) | 10 | - | - |
| CLTS | 142 (48%) | 145 (46%) | -19 | - | - |
| Control | 231 (49%) | 388 (61%) | 7 | - | - |
| Note: RRR = relative risk ratio; aRRR = adjusted relative risk ratio; CI = confidence interval; CLTS = community-led total sanitation; CG = care group model. |  |  |  |  |  |

To further explore the large number of hand rinsing events recorded, we utilised multinomial logistic regression models to calculate adjusted Relative Risk Ratios (aRRR) for a three-level handwashing outcome: no action, hand rinsing with water only, or handwashing with soap. In this analysis, not included in our original protocol, both interventions were associated with increased hand rinsing (i.e., handwashing with water only). Participants in the CLTS group were 1.8 times as likely to hand rinse compared to no hand hygiene action when compared to the control group (aRRR = 1.76, CI = 1.10, 2.83). Participants in the CLTS+CG group were 2.8 times as likely to hand rinse (aRRR = 2.80, CI = 1.81, 4.33) compared to no hand hygiene action (Table 3). Compared to the CLTS intervention, the CLTS+CG was associated with a slight increase in hand rinsing in the adjusted model (aRRR = 1.65, CI = 1.01, 2.69) (Table S3).

### Safe child faeces disposal

We observed 74 child faeces disposal opportunimes across all intervenmon groups at baseline and 122 opportunimes at endline (Figure 4). We found no evidence of differences in the odds of safe child faeces disposal among households in the CLTS group (aOR = 0.28; CI = 0.04, 1.94) and CLTS+CG group (aOR = 1.29; CI = 0.29, 5.71) compared to households in the control group eleven months after the intervention (Table S1). We also found no evidence of difference in the odds of safe child faeces disposal between the CLTS+CG group and CLTS group (aOR = 6.41; CI = 0.66, 62.33) (Table S2).

**Figure 4.**
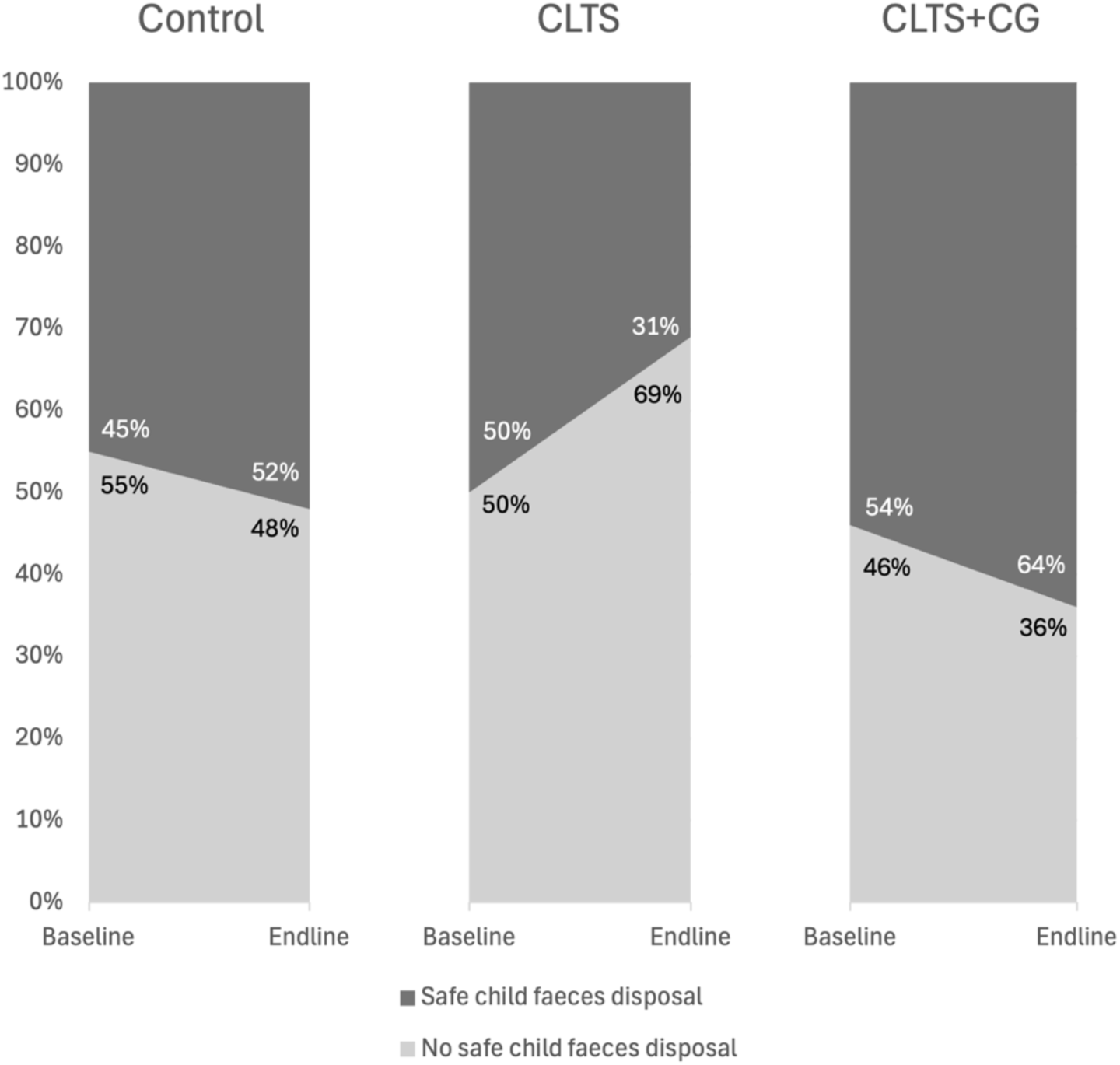
Observed child faeces disposal at baseline and endline in each trial arm.

## Discussion

This CBA trial investigated whether two district-wide community-based programmes promoting hygiene and sanitation behaviours were effective at improving observed handwashing and safe child faeces disposal in a rural area of Malawi. We found that the interventions – individually and in combination – did not increase handwashing with soap. These results are similar to a study in Nigeria that found that neither a standard CLTS intervention nor a CLTS intervention combined with a novel handwashing promotion intervention resulted in significant changes in handwashing with soap at key times^15^. However, both interventions in this study increased hand rinsing. Improvements in hand rinsing in both intervention groups suggest that current hygiene promotion interventions were effective at changing hand hygiene behaviour, but the promotion of soap for handwashing remains important for HHWS behaviour^42^. Although handwashing with water alone can substantially reduce the presence of bacteria on hands compared to no handwashing at all, handwashing with soap is more effective in removing bacteria from hands than handwashing with water only^42^, especially after faecal contact^43^.

Across the intervention groups, few households had soap available at the handwashing facility for handwashing with soap at both baseline and endline, suggesting that the availability of soap in the household is likely a barrier for effective hand hygiene in this setting. Similarly, in our main trial, only half of participants had soap of any kind (e.g., bar soap, liquid soap, or detergent) available in the home at the time of baseline data collection^31^.

Drawing on the capability, opportunity, and motivation (COM-B) framework for behaviour change^44^, the availability of soap represents an opportunity barrier for handwashing with soap^45^. An integrative review that found that the real or perceived availability of handwashing materials, such as soap, is associated with lower rates of handwashing with soap^46^. Other studies have identified the cost of soap as a barrier to hand hygiene ^46^, and affordability of soap may explain the low rates of handwashing with soap in this study. In a similar setting in rural Malawi, a study found that the high cost of soap was a barrier to consistent handwashing practices among new mothers and their guardians in healthcare facilities^47^. More research is needed to determine barriers to soap availability in the home and how future interventions can address these barriers in this setting. Soap is also often considered a luxury item and may be sequestered to keep it clean, prevent animals from consuming it, and stop children from playing with it^48^, limiting its availability for handwashing with soap at critical junctures^46,48,49^. Competing uses of soap in the household, such as the multiuse of soap for handwashing and laundry, may also lead to household members rationing soap and prioritising it for other uses^46^. Future research should additionally focus on soap use in the household, specifically to understand the prioritisation of soap in the household and how this may impact its use for handwashing.

The CLTS+CG intervention was slightly more effective than the CLTS intervention at increasing hand rinsing, suggesting that the CG model offers a promising additional channel to CLTS for changing hygiene behaviour. Previous studies in Malawi saw an increase in handwashing with soap at critical junctures as a result of a focussed behaviour change which utilised the CG model with integrated household visits^50,51^. In western Kenya^52^ and Zimbabwe^29^, the CG model increased the presence of a functional handwashing facility, a proxy measure for handwashing with soap, and handwashing with soap at critical junctures, respectively. In this study, smaller than anticipated changes in handwashing with soap may be due to how the CG model was implemented, in addition to limited soap availability. A process evaluation of the main trial associated with this sub-study found that exposure to household visits from CG members significantly increased the likelihood of having a handwashing facility^30^, but that CGs were primarily involved in promoting sanitation construction and use. Especially in households where handwashing soap is available, future hygiene interventions delivered using the CG model may consider focusing messages on promoting soap use.

Both interventions were not associated with any increase in safe child faeces disposal, suggesting that the promotion messages were insufficient to change behaviour. These findings align with a systematic review that found that most interventions targeting child faeces disposal did not result in significant improvements^18^. Similar to this study, almost all studies in the review included child faeces management as a component of a wider WASH intervention. In addition, most households had access to a latrine at endline, showing that the presence of a sanitation facility alone is not sufficient to ensure safe child faeces disposal^53^. Future behaviour change interventions may consider specifically targeting safe child faeces disposal to potentially achieve changes in behavioural and health outcomes^6,18^, especially as unsafe child faeces disposal was relatively high in all three groups at endline. In Malawi and some other countries, safe child faeces disposal is a requirement for achieving ODF status^19,32^, though CLTS interventions do not always target child faeces management in triggering activities to end open defecation^6,18^. Future CLTS interventions and government-led promotion campaigns, plus adaptations (e.g., CG approach), may consider additional or alternative messages for encouraging safe child faeces disposal^6^.

This study has several limitations. First, there were a limited number of child faeces disposal observations at both baseline and endline (74 and 122 observations, respectively) across all three groups. This sub-study was powered for sanitation outcomes in the main trial, so results from this analysis should be interpreted with caution. Second, data are based on observations which could lead to reactivity due to the Hawthorne effect (e.g., those being observed may modify their behaviour in response to being observed)^54^. To minimise this, enumerators mentioned they were generally interested in observing household behaviours when they visited households to avoid disclosing the behaviours of interest to the participants. Third, blinding of the intervention to participating households was not possible. However, the enumerators conducting the observations were blinded to the treatment allocation.

## Conclusion

The CLTS+CG and CLTS interventions were effective at increasing hand rinsing but not handwashing with soap. The availability of soap in the household and competing priorities for soap may be a barrier to handwashing in this setting, which warrants further research. The CLTS+CG intervention was slightly more effective than the CLTS intervention at increasing hand rinsing, suggesting that the Care Group model is a potentially promising additional behaviour change intervention. The interventions were not effective at increasing safe child faeces disposal, though the sample size was small. Future research can investigate the further integration of child faeces management into CLTS and behaviour change interventions at-scale.

## Acknowledgements

We would like to extend our gratitude to all the participating households who allowed us into their homes and made this study possible. Thank you also to the enumerators for their hard work collecting the data.

## Data availability

All data will be made publicly available in the LSHTM data repository.

## Code availability

Code to reproduce results will be made available via the LSHTM data repository.

## Supplementary information

**Table S1.** CLTS and CLTS+CG vs control for observed hand hygiene and safe child faeces disposal behaviours.

|  | Baseline, n (%) | Endline, n (%) | Difference, % | OR (95% CI) | aOR (95% CI) <sup>a</sup> | ICC |
| --- | --- | --- | --- | --- | --- | --- |
| **Handwashing with soap and water** |  |  |  |  |  | 0.120 |
| CLTS+CG | 17 (5%) | 12 (3%) | -2 | 1.42 (0.42, 4.77) | 1.87 (0.54, 6.40) |  |
| CLTS | 7 (2%) | 3 (1%) | -1 | 0.92 (0.16, 5.50) | 0.75 (0.13, 4.33) |  |
| Control | 12 (3%) | 7 (1%) | -2 | 1.00 (Ref) | 1.00 (Ref) |  |
| **Safe child faeces disposal** |  |  |  |  |  | 2.00e-36 |
| CLTS+CG | 14 (54%) | 23 (64%) | 10 | 1.16 (0.28, 4.79) | 1.29 (0.29, 5.71) |  |
| CLTS | 9 (50%) | 8 (31%) | -19 | 0.30 (0.06, 1.64) | 0.28 (0.04, 1.94) |  |
| Control | 13 (45%) | 31 (52%) | 7 | 1.00 (Ref) | 1.00 (Ref) |  |
| Note: OR = odds ratio; aOR = adjusted odds ratio; CI = confidence interval; ICC = intraclass correlation coefficient; CLTS = community-led total sanitation; CG = care group model. |  |  |  |  |  |  |
| <sup>a</sup> adjusted for community size, number of household members, at least one household member with a disability, wealth quintile, water source on plot, and village size. |  |  |  |  |  |  |

**Table S2.** CLTS+CG vs. CLTS for observed hand hygiene and safe child faeces disposal behaviours.

|  | Baseline, n (%) | Endline, n (%) | Difference, % | OR (95% CI) | aOR (95% CI) <sup>a</sup> |
| --- | --- | --- | --- | --- | --- |
| **Handwashing with soap and water** |  |  |  |  |  |
| CLTS+CG | 17 (5%) | 12 (3%) | -2 | 1.56 (0.25, 9.76) | 2.78 (0.49, 15.64) |
| CLTS | 7 (2%) | 3 (1%) | -1 | 1.00 (Ref) | 1.00 (Ref) |
| **Safe child faeces disposal** |  |  |  |  |  |
| CLTS+CG | 14 (54%) | 23 (64%) | 10 | 4.37 (0.60, 31.87) | 6.41 (0.66, 62.33) |
| CLTS | 9 (50%) | 8 (31%) | -19 | 1.00 (Ref) | 1.00 (Ref) |
| Note: OR = odds ratio; aOR = adjusted odds ratio; CI = confidence interval; ICC = intraclass correlation coefficient; CLTS = community-led total sanitation; CG = care group model. |  |  |  |  |  |
| <sup>a</sup> adjusted for community size, number of household members, at least one household member with a disability, wealth quintile, water source on plot, and village size. |  |  |  |  |  |

**Table S3.** CLTS+CG vs. CLTS for the three-level handwashing outcome variable.

|  | Baseline, n (%) | Endline, n (%) | Difference, % | RRR (95% CI) | aRRR (95% CI) <sup>a</sup> |
| --- | --- | --- | --- | --- | --- |
| **Handwashing with soap and water** |  |  |  |  |  |
| CLTS+CG | 17 (5%) | 12 (3%) | -2 | 1.89 (0.29, 12.32) | 3.55 (0.60, 20.79) |
| CLTS | 7 (2%) | 3 (1%) | -1 | 1.00 (Ref) | 1.00 (Ref) |
| **Hand rinsing** |  |  |  |  |  |
| CLTS+CG | 145 (44%) | 193 (55%) | 11 | 1.44 (0.88, 2.34) | 1.65 (1.01, 2.69) |
| CLTS | 153 (52%) | 172 (54%) | 2 | 1.00 (Ref) | 1.00 (Ref) |
| **No action** |  |  |  |  |  |
| CLTS+CG | 14 (54%) | 23 (64%) | 10 | (base) | (base) |
| CLTS | 9 (50%) | 8 (31%) | -19 | (base) | (base) |
| Note: RRR = relative risk ratio; aRRR = adjusted relative risk ratio; CI = confidence interval; CLTS = community-led total sanitation; CG = care group model. |  |  |  |  |  |
| <sup>a</sup> adjusted for community size, number of household members, at least one household member with a disability, wealth quintile, water source on plot, and village size. |  |  |  |  |  |

**Table S4.**
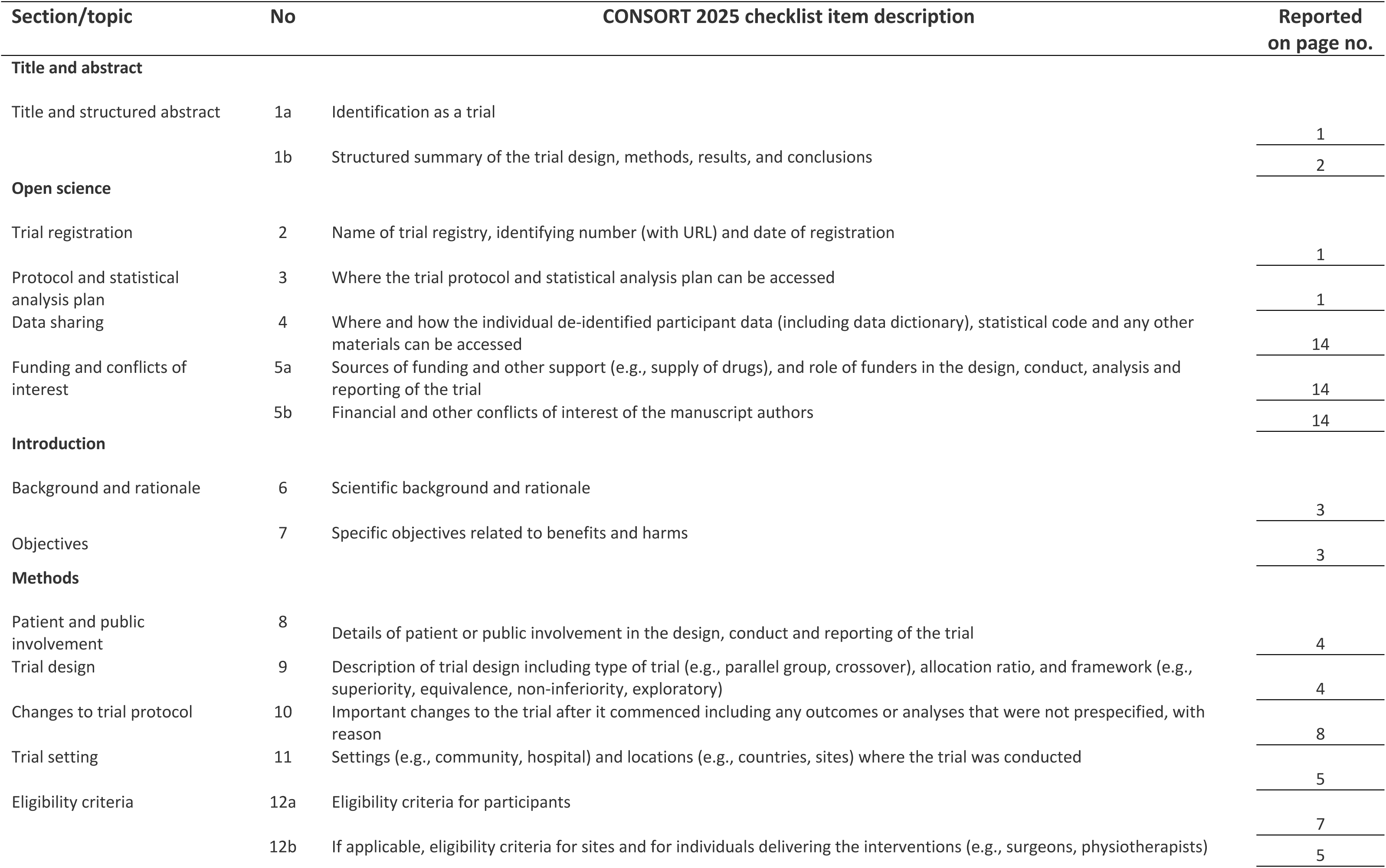
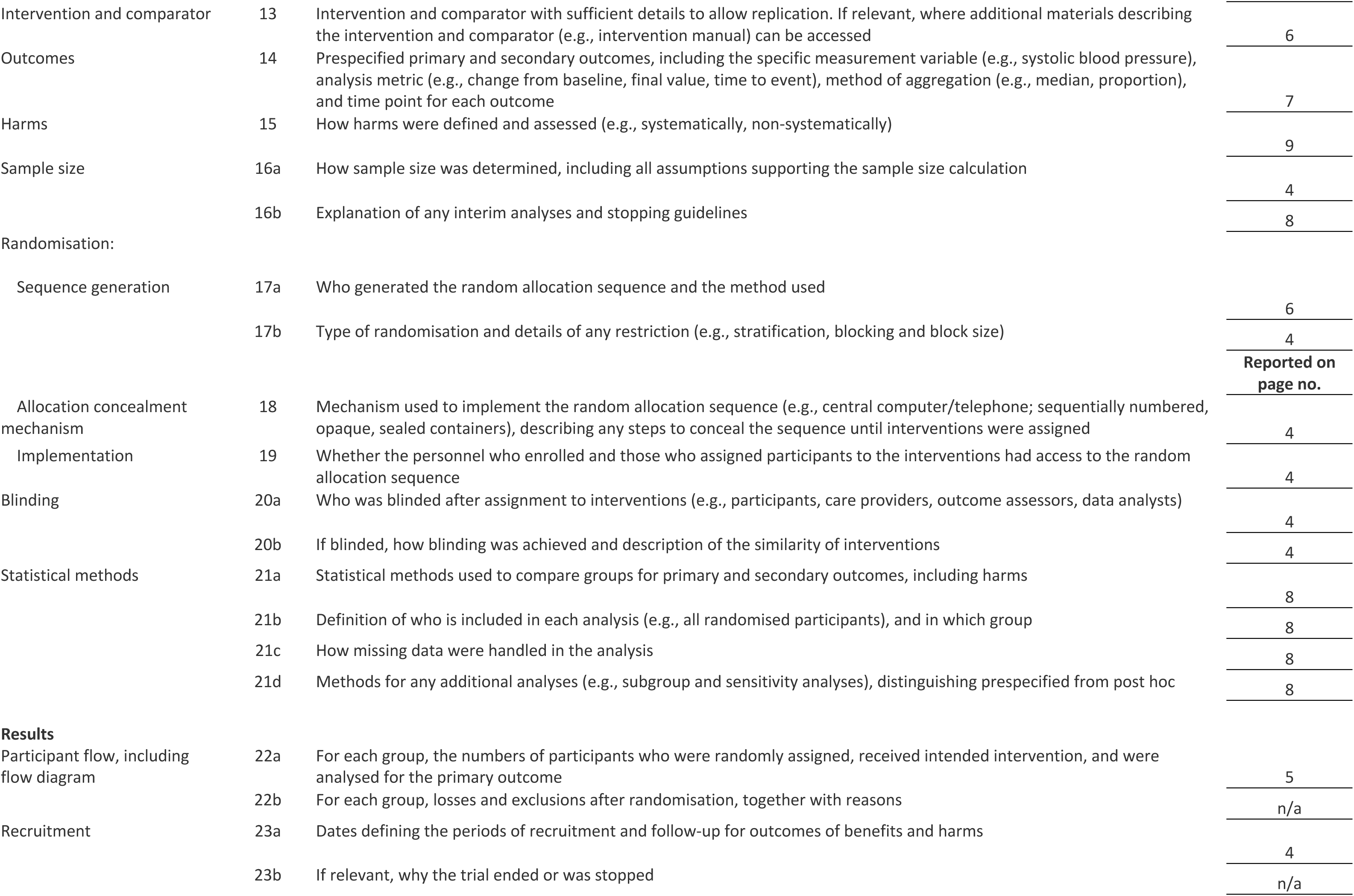
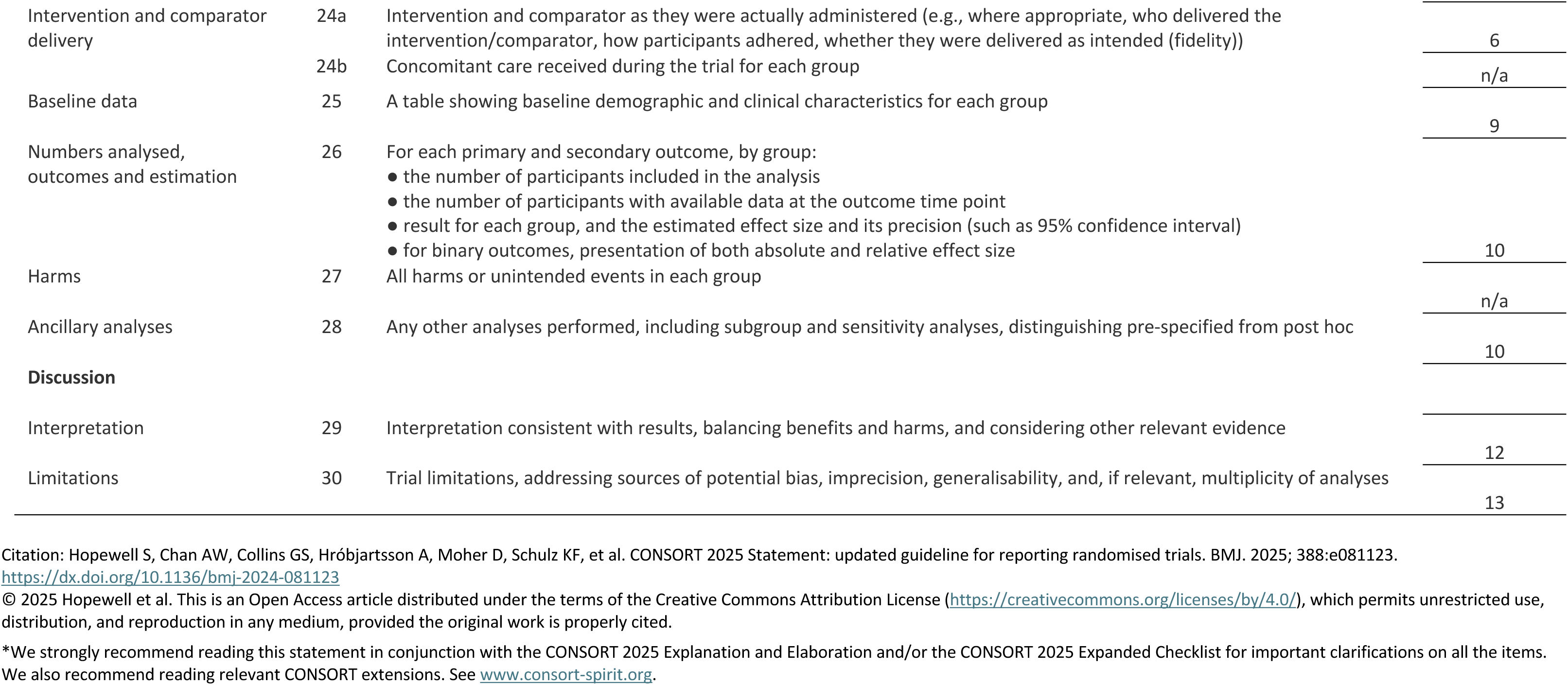
CONSORT checklist.

## References

(1) Wolf, J.; Johnston, R. B.; Ambelu, A.; Arnold, B. F.; Bain, R.; Brauer, M.; Brown, J.; Caruso, B. A.; Clasen, T.; Colford, J. M.; Mills, J. E.; Evans, B.; Freeman, M. C.; Gordon, B.; Kang, G.; Lanata, C. F.; Medlicott, K. O.; Prüss-Ustün, A.; Troeger, C.; Boisson, S.; Cumming, O. Burden of Disease Attributable to Unsafe Drinking Water, Sanitation, and Hygiene in Domestic Settings: A Global Analysis for Selected Adverse Health Outcomes. The Lancet 2023, 401 (10393), 2060–2071. 10.1016/S0140-6736(23)00458-0.

(2) Caruso, B. A.; Conrad, A.; Patrick, M.; Owens, A.; Kviten, K.; Zarella, O.; Rogers, H.; Sinharoy, S. S. Water, Sanitation, and Women’s Empowerment: A Systematic Review and Qualitative Metasynthesis. PLOS Water 2022, 1 (6), e0000026. 10.1371/journal.pwat.0000026.

(3) Ross, I.; Greco, G.; Adriano, Z.; Nala, R.; Brown, J.; Opondo, C.; Cumming, O. Impact of a Sanitation Intervention on Quality of Life and Mental Well-Being in Low-Income Urban Neighbourhoods of Maputo, Mozambique: An Observational Study. BMJ Open 2022, 12 (10), e062517. 10.1136/bmjopen-2022-062517.

(4) Sclar, G. D.; Garn, J. V.; Penakalapati, G.; Alexander, K. T.; Krauss, J.; Freeman, M. C.; Boisson, S.; Medlicott, K. O.; Clasen, T. Effects of Sanitation on Cognitive Development and School Absence: A Systematic Review. Int J Hyg Environ Health 2017, 220 (6), 917–927. 10.1016/j.ijheh.2017.06.010.

(5) Bauza, V.; Ye, W.; Liao, J.; Majorin, F.; Clasen, T. Interventions to Improve Sanitation for Preventing Diarrhoea. Cochrane Database of Systematic Reviews 2023, 2023 (1). 10.1002/14651858.CD013328.pub2.

(6) Majorin, F.; Torondel, B.; Ka Seen Chan, G.; Clasen, T. Interventions to Improve Disposal of Child Faeces for Preventing Diarrhoea and Soil-Transmitted Helminth Infection. Cochrane Database of Systematic Reviews 2019, 2019 (9). 10.1002/14651858.CD011055.pub2.

(7) Wolf, J.; Hubbard, S.; Brauer, M.; Ambelu, A.; Arnold, B. F.; Bain, R.; Bauza, V.; Brown, J.; Caruso, B. A.; Clasen, T.; Colford, J. M.; Freeman, M. C.; Gordon, B.; Johnston, R. B.; Mertens, A.; Prüss-Ustün, A.; Ross, I.; Stanaway, J.; Zhao, J. T.; Cumming, O.; Boisson, S. Effectiveness of Interventions to Improve Drinking Water, Sanitation, and Handwashing with Soap on Risk of Diarrhoeal Disease in Children in Low-Income and Middle-Income Settings: A Systematic Review and Meta-Analysis. The Lancet 2022, 400 (10345), 48–59. 10.1016/S0140-6736(22)00937-0.

(8) Ross, I.; Bick, S.; Ayieko, P.; Dreibelbis, R.; Wolf, J.; Freeman, M. C.; Allen, E.; Brauer, M.; Cumming, O. Effectiveness of Handwashing with Soap for Preventing Acute Respiratory Infections in Low-Income and Middle-Income Countries: A Systematic Review and Meta-Analysis. The Lancet 2023, 401 (10389), 1681–1690. 10.1016/S0140-6736(23)00021-1.

(9) Strunz, E. C.; Addiss, D. G.; Stocks, M. E.; Ogden, S.; Utzinger, J.; Freeman, M. C. Water, Sanitation, Hygiene, and Soil-Transmitted Helminth Infection: A Systematic Review and Meta-Analysis. PLoS Med 2014, 11 (3), e1001620. 10.1371/journal.pmed.1001620.

(10) Stocks, M. E.; Ogden, S.; Haddad, D.; Addiss, D. G.; McGuire, C.; Freeman, M. C. Effect of Water, Sanitation, and Hygiene on the Prevention of Trachoma: A Systematic Review and Meta-Analysis. PLoS Med 2014, 11 (2), e1001605. 10.1371/journal.pmed.1001605.

(11) De Buck, E.; Van Remoortel, H.; Hannes, K.; Govender, T.; Naidoo, S.; Avau, B.; Vande Veegaete, A.; Musekiwa, A.; Lutje, V.; Cargo, M.; Mosler, H.-J.; Vandekerckhove, P.; Young, T. 10.1016/j.Ijheh.2025.114519; 2017.

(12) MacLeod, C.; Davies, K.; Mwenge, M. M.; Chipungu, J.; Cumming, O.; Dreibelbis, R. Behaviour Change Interventions to Improve Household Sanitation and Hygiene Practices in Urban Settings: A Systematic Scoping Review. Int J Hyg Environ Health 2025, 264, 114519. 10.1016/j.ijheh.2025.114519.

(13) Kar, K.; Chambers, R. Handbook on Community Led Total Sanitation; 2008.

(14) Venkataramanan, V.; Crocker, J.; Karon, A.; Bartram, J. Community-Led Total Sanitation: A Mixed-Methods Systematic Review of Evidence and Its Quality. Environ Health Perspect 2018, 126 (2). 10.1289/EHP1965.

(15) Biran, A.; White, S.; Awe, B.; Greenland, K.; Akabike, K.; Chuktu, N.; Aunger, R.; Curtis, V.; Schmidt, W.; Van der Voorden, C. A Cluster-Randomised Trial to Evaluate an Intervention to Promote Handwashing in Rural Nigeria. Int J Environ Health Res 2022, 32 (3), 579–594. 10.1080/09603123.2020.1788712.

(16) Yeboah-Antwi, K.; MacLeod, W. B.; Biemba, G.; Sijenyi, P.; Höhne, A.; Verstraete, L.; McCallum, C. M.; Hamer, D. H. Improving Sanitation and Hygiene through Community-Led Total Sanitation: The Zambian Experience. Am J Trop Med Hyg 2019, 100 (4), 1005–1012. 10.4269/ajtmh.18-0632.

(17) Briceño, B.; Coville, A.; Gertler, P.; Martinez, S. Are There Synergies from Combining Hygiene and Sanitation Promotion Campaigns: Evidence from a Large-Scale Cluster-Randomized Trial in Rural Tanzania. PLoS One 2017, 12 (11), e0186228. 10.1371/journal.pone.0186228.

(18) Morita, T.; Godfrey, S.; George, C. M. Systematic Review of Evidence on the Effectiveness of Safe Child Faeces Disposal Interventions. Tropical Medicine & International Health 2016, 21 (11), 1403–1419. 10.1111/tmi.12773.

(19) Thomas, A.; Bevan, J. Developing and Monitoring Protocol for the Elimination of Open Defecation in Sub-Saharan Africa; 2013.

(20) Morse, T.; Chidziwisano, K.; Tilley, E.; Malolo, R.; Kumwenda, S.; Musaya, J.; Cairncross, S. Developing a Contextually Appropriate Integrated Hygiene Intervention to Achieve Sustained Reductions in Diarrheal Diseases. Sustainability 2019, 11 (17), 4656. 10.3390/su11174656.

(21) Core Group; USAID; Relief W; Food for the Hungry. Care Group Results: Implementor’s List; 2021.

(22) Pieterse, P.; Walsh, A.; Chirwa, E.; Chikalipo, M.; Msowoya, C.; Mambulasa, J.; Matthews, A. Evaluating the Role That Care Groups Play in Providing Breastfeeding and Infant Feeding Support at Community Level: A Qualitative Study in Dedza District in Malawi. HRB Open Res 2023, 6, 44. 10.12688/hrbopenres.13736.1.

(23) Gladstone, M. J.; Chandna, J.; Kandawasvika, G.; Ntozini, R.; Majo, F. D.; Tavengwa, N. V.; Mbuya, M. N. N.; Mangwadu, G. T.; Chigumira, A.; Chasokela, C. M.; Moulton, L. H.; Stoltzfus, R. J.; Humphrey, J. H.; Prendergast, A. J. Independent and Combined Effects of Improved Water, Sanitation, and Hygiene (WASH) and Improved Complementary Feeding on Early Neurodevelopment among Children Born to HIV-Negative Mothers in Rural Zimbabwe: Substudy of a Cluster-Randomized Trial. PLoS Med 2019, 16 (3), e1002766. 10.1371/journal.pmed.1002766.

(24) Perry, H.; Morrow, M.; Borger, S.; Weiss, J.; DeCoster, M.; Davis, T.; Ernst, P. Care Groups I: An Innovative Community-Based Strategy for Improving Maternal, Neonatal, and Child Health in Resource-Constrained Settings. Glob Health Sci Pract 2015, 3 (3), 358–369. 10.9745/GHSP-D-15-00051.

(25) Panulo, M.; Chidziwisano, K.; Beattie, T. K.; Tilley, E.; Kambala, C.; Morse, T. Process Evaluation of “The Hygienic Family” Intervention: A Community-Based Water, Sanitation, and Hygiene Project in Rural Malawi. Int J Environ Res Public Health 2022, 19 (11), 6771. 10.3390/ijerph19116771.

(26) Perry, H.; Morrow, M.; Davis, T.; Borger, S.; Weiss, J.; DeCoster, M.; Ernst, P. Care Groups - an Effective-Community-Based Delivery Strategy for Improving Reproductive, Maternal, Neonatal and Child Health in High-Mortality, Resource-Constrained Settings (a Guide for Policy-Makers and Donors); 2014.

(27) Pieterse, P.; Matthews, A.; Walsh, A.; Chirwa, E. Exploring How and Why Care Groups Work to Improve Infant Feeding Practices in Low- and Middle-Income Countries: A Realist Review Protocol. Syst Rev 2020, 9 (1), 237. 10.1186/s13643-020-01497-1.

(28) Freeman, M. C.; Ellis, A. S.; Ogutu, E. A.; Caruso, B. A.; Linabarger, M.; Micek, K.; Muga, R.; Girard, A. W.; Wodnik, B. K.; Jacob Arriola, K. Impact of a Demand-Side Integrated WASH and Nutrition Community-Based Care Group Intervention on Behavioural Change: A Randomised Controlled Trial in Western Kenya. BMJ Glob Health 2020, 5 (11), e002806. 10.1136/bmjgh-2020-002806.

(29) Matsungo, T. M.; Kamazizwa, F.; Mavhudzi, T.; Makota, S.; Kamunda, B.; Matsinde, C.; Chagwena, D.; Mukudoka, K.; Chopera, P. Influence of Care Group Participation on Infant and Young Child Feeding, Dietary Diversity, WASH Behaviours and Nutrition Outcomes in Rural Zimbabwe. BMJ Nutr Prev Health 2023, 6 (2), 164–172. 10.1136/bmjnph-2023-000627.

(30) Panulo, M.; Chidziwisano, K.; MacLeod, C.; Kapazga, T.; Dreibelbis, R.; Beattie, T. K.; Morse, T. CLTS Implementation in Malawi: Process Evaluation of a Sanitation and Hygiene Intervention. 2024. 10.31219/osf.io/9bfqd.

(31) Chidziwisano, K.; Panulo, M.; MacLeod, C.; Vigneri, M.; White, B.; Wells, J.; Ross, I.; Morse, T.; Dreibelbis, R. Water, Sanitation, and Hygiene for Everyone Intervention Study: Protocol for a Controlled Before-and-After Trial. JMIR Res Protoc 2025, 14, e68280. 10.2196/68280.

(32) Malawi Ministry of Health and Population. National Sanitation and Hygiene Strategy; Lilongwe, Malawi, 2018.

(33) Chidziwisano, K.; Tilley, E.; Malolo, R.; Kumwenda, S.; Musaya, J.; Morse, T. Risk Factors Associated with Feeding Children under 2 Years in Rural Malawi—A Formative Study. Int J Environ Res Public Health 2019, 16 (12), 2146. 10.3390/ijerph16122146.

(34) WHO/UNICEF Joint Monitoring Programme for Water Supply, S. and H. Sanitation. WHO/UNICEF Joint Monitoring Programme for Water Supply, Sanitation and Hygiene.

(35) R Core Team. R: A Language and Environment for Statistical Computing. R Foundation for Statistical Computing: Vienna, Austria 2021.

(36) Hollis, S.; Campbell, F. What Is Meant by Intention to Treat Analysis? Survey of Published Randomised Controlled Trials. BMJ 1999, 319 (7211), 670–674. 10.1136/bmj.319.7211.670.

(37) Vyas, S.; Kumaranayake, L. Constructing Socio-Economic Status Indices: How to Use Principal Components Analysis. Health Policy Plan 2006, 21 (6), 459–468. 10.1093/heapol/czl029.

(38) Hubbard, A. E.; Ahern, J.; Fleischer, N. L.; Laan, M. Van der; Lippman, S. A.; Jewell, N.; Bruckner, T.; Satariano, W. A. To GEE or Not to GEE. Epidemiology 2010, 21 (4), 467–474. 10.1097/EDE.0b013e3181caeb90.

(39) Bottomley, C.; Kirby, M. J.; Lindsay, S. W.; Alexander, N. Can the Buck Always Be Passed to the Highest Level of Clustering? BMC Med Res Methodol 2016, 16 (1), 29. 10.1186/s12874-016-0127-1.

(40) Hayes, R.; Moulton, L. Cluster Randomized Trials. Taylor & Francis Group: Boca Raton, FL 2009.

(41) Washington Group on Disability Statistics. The Washington Group Short Set on Functioning (WG-SS); Hyattsville, Maryland, 2022.

(42) Burton, M.; Cobb, E.; Donachie, P.; Judah, G.; Curtis, V.; Schmidt, W.-P. The Effect of Handwashing with Water or Soap on Bacterial Contamination of Hands. Int J Environ Res Public Health 2011, 8 (1), 97–104. 10.3390/ijerph8010097.

(43) Wolf, J.; Johnston, R.; Freeman, M. C.; Ram, P. K.; Slaymaker, T.; Laurenz, E.; Prüss-Ustün, A. Handwashing with Soap after Potential Faecal Contact: Global, Regional and Country Estimates. Int J Epidemiol 2019, 48 (4), 1204–1218. 10.1093/ije/dyy253.

(44) Michie, S.; van Stralen, M. M.; West, R. The Behaviour Change Wheel: A New Method for Characterising and Designing Behaviour Change Interventions. Implementation Science 2011, 6 (1), 42. 10.1186/1748-5908-6-42.

(45) Caruso, B. A.; Snyder, J. S.; O’Brien, L. A.; LaFon, E.; Files, K.; Shoaib, D. M.; Prasad, S. K.; Rogers, H.; Cumming, O.; Esteves Mills, J.; Gordon, B.; Wolfe, M. K.; Freeman, M. C. Behavioural Factors Influencing Hand Hygiene Practices across Domestic, Institutional, and Public Community Settings: A Systematic Review. March 2025. 10.1101/2025.03.11.25323561.

(46) Ezezika, O.; Heng, J.; Fatima, K.; Mohamed, A.; Barrett, K. What Are the Barriers and Facilitators to Community Handwashing with Water and Soap? A Systematic Review. PLOS Global Public Health 2023, 3 (4), e0001720. 10.1371/journal.pgph.0001720.

(47) Luwe, K.; Chidziwisano, K.; Davies, K.; Morse, T.; Dreibelbis, R. Handwashing Practices among New Mothers and Their Guardians: A Mixed-Methods Observational Study in Healthcare Facilities and Households in Rural Malawi. October 2024. 10.1101/2024.10.11.24315284.

(48) Bowen, A.; Agboatwalla, M.; Ayers, T.; Tobery, T.; Tariq, M.; Luby, S. P. Sustained Improvements in Handwashing Indicators More than 5 Years after a Cluster-randomised, Community-based Trial of Handwashing Promotion in Karachi, Pakistan. Tropical Medicine & International Health 2013, 18 (3), 259–267. 10.1111/tmi.12046.

(49) Biran, A.; Schmidt, W.; Zeleke, L.; Emukule, H.; Khay, H.; Parker, J.; Peprah, D. Hygiene and Sanitation Practices amongst Residents of Three Long-term Refugee Camps in Thailand, Ethiopia and Kenya. Tropical Medicine & International Health 2012, 17 (9), 1133–1141. 10.1111/j.1365-3156.2012.03045.x.

(50) Morse, T.; Tilley, E.; Chidziwisano, K.; Malolo, R.; Musaya, J. Health Outcomes of an Integrated Behaviour-Centred Water, Sanitation, Hygiene and Food Safety Intervention–A Randomised before and after Trial. Int J Environ Res Public Health 2020, 17 (8), 2648. 10.3390/ijerph17082648.

(51) Chidziwisano, K.; Tilley, E.; Morse, T. Self-Reported Versus Observed Measures: Validation of Child Caregiver Food Hygiene Practices in Rural Malawi. Int J Environ Res Public Health 2020, 17 (12), 4498. 10.3390/ijerph17124498.

(52) Freeman, M. C.; Ellis, A. S.; Ogutu, E. A.; Caruso, B. A.; Linabarger, M.; Micek, K.; Muga, R.; Girard, A. W.; Wodnik, B. K.; Jacob Arriola, K. Impact of a Demand-Side Integrated WASH and Nutrition Community-Based Care Group Intervention on Behavioural Change: A Randomised Controlled Trial in Western Kenya. BMJ Glob Health 2020, 5 (11), e002806. 10.1136/bmjgh-2020-002806.

(53) Mugel, S. G.; Clasen, T. F.; Bauza, V. Global Practices, Geographic Variation, and Determinants of Child Feces Disposal in 42 Low- and Middle-Income Countries: An Analysis of Standardized Cross-Sectional National Surveys from 2016 – 2020. Int J Hyg Environ Health 2022, 245, 114024. 10.1016/j.ijheh.2022.114024.

(54) Sedgwick, P.; Greenwood, N. Understanding the Hawthorne Effect. BMJ 2015, h4672. 10.1136/bmj.h4672.

